# First evidence of zoonotic spillover of MERS-CoV into occupationally exposed populations in Somalia

**DOI:** 10.64898/2026.03.17.26348312

**Authors:** Marian Warsame, Jamila Aden, Alice Simniceanu, Mutawe Lubogo, Samuel MS Cheng, Mahamoud Mohamed Hussein, Sahra Issa Mohamed, Abdirahman Osman Abdikadir, Abdiwali Mohamed Ahmed, Abdirahman Yasin Ahmed, Ahmed Mohamud Ahmed, Abduljalil Abdullahi Ali, Adnan Mohamed Ali, Omar Abdikarin Ali, Ali Mohamed Arif, Abdirashid Abdulkadir Bujeti, Abdirahman Jama Farah, Abdihakim M Hassan Hanafi, Abdulkadir Mohamed Hassan, Mohamud Abshir Hassan, Muse Ahmed Hassan, Jenson CH Ho, Fatima Abdullahi Hussein, Hassan Ahmed Hussein, Bishar Abdi Jama, John KC Li, Mohamed Adan Mohamed, Mustafe Abdirizak Mohamoud, Omar Mohamed Mohamud, Hala Abou El Naja, Mohamed Bashir Nuur, Omar Ali Said, Abdinur Abdullahi Salad, Abdullah Al Sayafi, Abukar Nor Farah Shurie, Maria D Van Kerkhove, Amal Barakat, Mohamd Mohamud Bidey, Malik Peiris, Ruth McCabe, Sophie von Dobschuetz

## Abstract

Dromedary camels are the main reservoir for Middle East respiratory syndrome coronavirus (MERS-CoV), a re-emerging infectious disease with pandemic potential. Somalia harbours approximately 32% of dromedary camels globally. We investigated current and past MERS-CoV infections among occupationally-exposed workers in slaughterhouses, dairy farms, livestock markets and a quarantine station. Sera and nasopharyngeal/oropharyngeal swabs from 770 workers were analysed for MERS-CoV antibodies by Enzyme-Linked Immunosorbent Assay (ELISA) and virus neutralization and for viral RNA by Real Star® MERS-CoV Reverse Transcription Polymerase Chain Reaction (RT-PCR). One farm worker with no travel history in the Qardo district, Karkar region, Puntland was sero-positive by ELISA and virus neutralization, providing the first-ever evidence of zoonotic spillover of MERS-CoV to humans in Somalia. This finding highlights the need to strengthen MERS-CoV surveillance across Somalia, along with an urgent need to strengthen national laboratory capacity and integrate MERS into diagnostic algorithms to generate accurate and reliable infection data and studies to understand the socio-cultural and potential risk factors for MERS-CoV.

## Introduction

The Middle East respiratory syndrome coronavirus (MERS-CoV) is a zoonotic virus with dromedary camels as the source of zoonotic infection (Reusken et al., 2013a; Reusken et al., 2013b; Haagmans et al., 2014; Azhan et al., 2014). The first human case of MERS-CoV was reported in Saudi Arabia in 2012 (Zaki et al., 2012). As of January 2026, 2,635 confirmed cases have been reported to the World Health Organization (WHO), most of them from Saudi Arabia. (WHO, 2025) Transmission of MERS-CoV from camels to humans is thought to occur through direct contact with the respiratory droplets or saliva of camels or through the consumption of camel products such as milk and undercooked camel meat (Reusken et al., 2014, Azhar et al., 2014, Sikkema et al., 2019, Alraddadi et al., 2016, Khudhair et al., 2019). Most cases of human-to-human transmission reported to date have occurred in healthcare facilities, sometimes leading to large outbreaks in Saudi Arabia, the United Arab Emirates and the Republic of Korea (Assiri et al., 2013, Oboho, Alhosani et al., 2016, Lee et al., 2017, Al-Abdallat et al., 2014, Memish et al., 2013, Alenazi et al. 2017). Human-to-human transmission has also sporadically occurred in households (Omrani et al., 2013) (Elkholy et al., 2020) (Memish et al., 2013) (Arwady et al., 2016). In healthcare settings, MERS-CoV can be transmitted between patients and healthcare workers, between patients or from patients to visitors (Hui et al., 2018), (Elkholy et al., 2020) (Alfaraj et al., 2018). MERS-CoV has been detected in clinical specimens such as sputum, endotracheal aspirate, bronchoalveolar lavage, nasal or nasopharyngeal swabs, urine, faeces, blood, and lung tissue from patients (Lee et al., 2017) (Fagbo et al., 2017). When investigating human exposure to MERS-CoV in Saudi Arabia, Muller et al., (2015) found a 15- and 23-fold higher seroprevalence of antibodies in camel herders and slaughterhouse workers, respectively, compared to the general population. They concluded that these individuals might also be the source of infection for cases with confirmed MERS but with no previous exposure to camels (WHO, 2026).

There is evidence of active circulation of MERS-CoV in dromedary camels in sub–Saharan Africa, including in Ethiopia and Kenya (Miguel et al., 2017), (Zhou et al., 2023) (Ommeh et al., 2018) (Kiambi et al., 2018), (Kiyong’a et al., 2020). In 2018 – 2020, an outbreak in camels in Kenya was detected, with reports of possible spillover into humans (Ngere et al., 2022). Multiple other studies have also detected MERS-CoV antibodies in camel handlers in Kenya (Liljander et al., 2016, Kiyong’s et al., 2020, Munyua et al., 2021), underlining the risk of infection from occupational exposure to dromedary camels beyond the Arabian Peninsula.

As of 2016, Somalia had more camels (7.1 million) than any country in the world, accounting for 43% of dromedary camels in Africa and 32% of dromedaries globally. Together with other livestock, camels constitute the backbone of the country’s economy, accounting for about 61% of gross domestic product and 79% of export earnings (Akester, 2018). Camels not only provide milk, meat and means of transportation but are also highly valued for their role in traditional social activities, for example, as the bride price (Yarad) or as blood money to the family of the deceased in clan feuds (Hussein, 2014). Despite this central role of camels in the culture and economy of Somalia, there is no strategic surveillance for MERS-CoV in humans in Somalia, nor have any complementary research studies been conducted to date to assess the burden of MERS in this country (Müller et al., 2014). In this first study of its type in Somalia, we tested 770 occupationally exposed humans in eight regions for MERS-CoV RNA and antibodies.

## Methods and materials

### Study areas and facilities

The study was conducted in the eight regions with the highest concentration of dromedary camels in Somalia: Bari, Karkar, Nugal, Mudug, Galgadud, Benadir, Bay and Lower Juba (Figure 1). In each region, the town with the highest concentration of camels was purposely selected.

**Figure 1.**
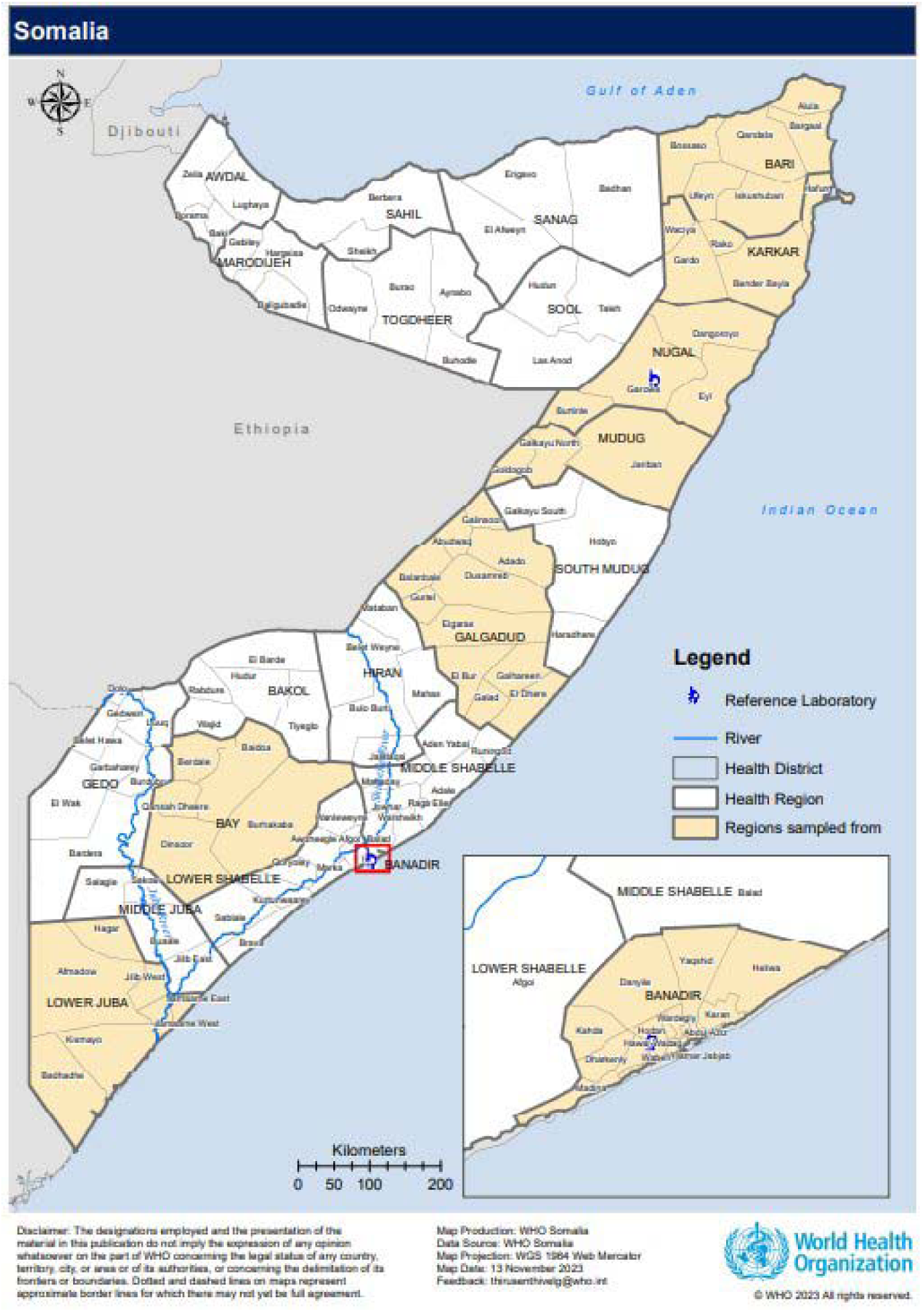
District level map of Somalia, with yellow shading highlighting the regions surveyed in this work.

Within these regions, camel markets, dairy farms, slaughterhouses and quarantine facilities were all considered high-risk for occupational exposure to MERS-CoV thus selected for inclusion in the study. Specific facilities were further selected based on their availability and location in each of the major cities, and whether they met the selection criteria, including contained camels with workers aged at least 18 years old. All camel markets trading live camels and slaughterhouses located in large cities, alongside dairy farms and quarantine facilities located in each region were contacted. All contacted facilities which gave consent were subsequently enrolled in the study.

Camel markets in all towns surveyed were similar in structure; being in an open area with separate sections for dromedary camels, goats, cows and sheep, with each owner keeping their herd together. The slaughterhouses were closed spaces with a wall (Kismayo, Garowe, Galkayo, Qardo and Bosaso) and were run by private individuals or groups. Camels were slaughtered in these facilities, and the meat sold to meat traders and restaurants for local consumption. The Somali Meat Company (SOMEAT) in Benadir, a modern private slaughterhouse processing meat for exporters, was also included in this study. All the dairy farms, except those in Mogadishu, Benadir, were located on the outskirts of the towns in open areas where the camels stayed from sunset to sunrise and let out to graze during the day. On the dairy farms in Mogadishu camels were kept and fed for as long as they provided milk and taken to grazing land until they could produce milk again. The quarantine facility in Bosaso city was strategically located near camel markets to effectively monitor and control the movement of camels, helping prevent the spread of diseases like MERS-CoV. This facility varies in structure ranging from open areas to enclosed spaces and is operated by local authorities. Key health measures implemented include vaccination for other animals such as sheep, goats, and cattle and rigorous health screenings, with stringent sanitation and hygiene protocols to ensure the facilities themselves do not become transmission hubs. The facility is equipped to conduct regular health checks and isolate new or suspected infected camels from healthy ones to prevent disease transmission to other animals or humans. Furthermore, the facility carries out laboratory testing for common infectious and zoonotic diseases such as brucellosis, where samples such as blood and nasal swabs are collected for diagnostic confirmation before animals are cleared for trade or transport.

### Study design and population

The WHO Unity Study protocol for cross-sectional seroepidemiologic studies of MERS-CoV infection in high-risk populations in contact with dromedary camels was used for this study (WHO, 2024). Study participants were selected from the enrolled facilities in the selected study localities if they were directly involved in handling live or deceased camels or camel products. This included specific roles according to the different facility types: camel owners and/or sellers, those purchasing camels, brokers (dalal) and cleaners within markets; those responsible for slaughtering, skinning, and evisceration of camels, cleaners, porters and guards within slaughterhouses; cleaners of the camel shelters, the handlers of the camel waste, birthing and milking personnel and guards within dairy farms; and those involved in daily animal health monitoring, animal handling (e.g. veterinarians, animal health assistants, livestock inspectors), cleaners, and security guards (responsible for collecting biological samples, vaccination, inspecting of the camels upon arrival, isolation processes, maintaining hygiene standards) within the quarantine centre.

Workers that met this eligibility criteria, in addition to being aged at least 18 years old and working in the facility at least six months prior to sampling, were invited to participate and provided with information about the study objectives and procedures. Those who consented were subsequently enrolled in the study. Ethical Clearance was approved from the East Africa University Research Ethical Review Committee.

### Data and sample collection

Data and samples were collected between 22 February and 9 April 2023.

*Interviews:* The structured questionnaire developed by WHO (WHO, 2024) for each type of study facility (livestock market, slaughterhouse, dairy farm, and quarantine station) was field tested and adapted based on feedback. After obtaining informed consent, information was collected on demographics (age, gender etc.), occupational exposure (type of job in the facility, duration of exposure), use of personal protective equipment (PPE) and hygiene practices (whether participants used PPE while working, the type of PPE worn, when hand hygiene performed), as well as exposure to camel products for food or medicine.

*Biological sampling:* A total of 5 ml of blood was collected in a tube without anticoagulant and immediately transported in a cool box from the collection site to an assigned laboratory facility where it was centrifuged (1500 × g, 15 minutes), sera separated and stored at −20 °C in the state reference laboratories.

Nasopharyngeal and oropharyngeal swabs were collected from each participant to test for the presence of MERS-CoV. The swabs were placed in viral medium transport, stored temporarily in a cool box until they were transported to the assigned laboratory at the study site.

Both sera (for serological analyses) and swabs (for virological analyses) were sent to the National Public Health Laboratory in Mogadishu for samples for Galgadud, Benadir, Bay and Lower Jubba regions and to the Puntland State Public Health Laboratory in Garowe for samples from Mudug, Bari, Karkar and Nugal regions.

### Laboratory analysis

*Virus RNA detection:* Nasopharyngeal and oropharyngeal swabs were screened for qualitative detection of MERS-CoV-specific viral RNA (evidence of active infection) using the Real Star® MERS-CoV RT-PCR Kit 1.0. This consists of two independent assays, one targeting a region upstream of the E gene (upE) and the other targeting the open reading frame 1a (orf1a) of the MERS-CoV genome. The PCR type used was Rotor-Gene® Q5 & 6 plex (QIAGEN) and the type of RNA extraction kit used was QIAamp® Viral RNA Mini Kit (QIAGEN).

*ELISA assay for MERS-CoV antibody:* The semi-quantitative anti-MERS-CoV IgG human ELISA (EUROIMMUN anti-MERS-CoV ELISA-IgG) was used to detect specific IgG antibodies against MERS-CoV in human sera indicative of previous infection. The test was performed according to the manufacturer’s instructions(Ag, 2025). The result was evaluated semi-quantitatively using ratio values as follows:

Ratio = Extinction value of the sample/the extinction value of the calibrator.

Serum samples with a ratio greater than 1.1 were considered positive for MERS-CoV antibodies. All borderline (defined by an OD ratio (test sample/calibrator) ≥ 0.8 and < 1.1) and positive samples, along with 73 randomly selected negative sera as controls, were sent to the reference laboratory at the University of Hong Kong for confirmation by a neutralization assay and investigation of potential ELISA cross-reactivity with SARS-CoV-2, given its widespread circulation.

*MERS-CoV Spike Pseudoparticle Neutralization Test (ppNT*): The ppNT assay was performed as previously described (Perera et al 2013) using 96-well tissue culture plates (TPP Techno Plastic Products AG, Trasadingen, Switzerland) in triplicate. Luciferase-expressing HIV/MERS pseudoparticles (5 ng of p24) were pre-incubated with diluted serum (1:10) at 4°C for 30 minutes. The serum-pseudoparticle mixtures were then added to pre-formed Vero E6 cell monolayers (STCC CRL-1586). After a 48-hour incubation period, the cells were lysed in 20ul lysis buffer and followed addition of 100ul of luciferase substrate (Promega Corporation, Madison, US). Luciferase activity was quantified using a Microbeta luminometer. A positive result was defined as ≥90% reduction of luciferase activity compared to control. Positive sera were titrated to endpoint to define the highest serum dilution that resulted in a ≥90% reduction in luciferase activity compared to control wells without antibodies, regarded as the ppNT90 neutralizing titre.

*MERS-CoV plaque reduction neutralization test (PRNT):* The PRNT was performed as previously described (Park et al 2015) in duplicate using 24-well tissue culture plates (TPP Techno Plastic Products AG, Trasadingen, Switzerland) in a biosafety level 3 facility using Vero E6 cells. Sera were heat-inactivated at 56 °C for 30 min prior to testing. Serially diluted serum samples (starting dilution 1:10) were incubated with 30–40 plaque-forming units of virus (EMC strain) for 1 hour at 37 °C and the virus–serum mix was added onto pre-formed Vero E6 cell monolayers and incubated for 1 hour at 37 °C in a 5% CO2 incubator. The virus-antibody inoculum was then discarded, and the cell monolayer was overlaid with 1% agarose in cell culture medium. After 3 days incubation, the plates were fixed with 10% formalin in PBS overnight, stained with 1% crystal violet in ethanol and the plaques were counted visually. Antibody titres were defined as the highest serum dilution that resulted in ≥ 50% (PRNT50) or ≥ 90% (PRNT90) reduction in the number of virus plaques compared to no-serum control wells. The average plaque numbers observed in the duplicate dilution-series was used for this computation. Virus back titrations, positive and negative control sera were included in every experiment.

*SARS-CoV-2 Anti-Spike-RBD ELISA (IgG):* The SARS-CoV-2 Anti-Spike-RBD ELISA (IgG) was conducted as previously described (Perera et al 2020). In brief, 96-well ELISA plates (Nunc MaxiSorp, Thermo Fisher Scientific) were coated overnight with 100 ng/well of purified recombinant RBD protein in PBS buffer. The plates were then blocked with 100 μL of Chonblock blocking buffer (Chondrex Inc, Redmond, US) per well and incubated at room temperature for 2 hours. Serum samples, diluted 1:100 in Chonblock blocking buffer, were tested in duplicate. These samples were added to the wells and incubated for 2 hours at 37°C. After extensive washing with PBS containing 0.1% Tween 20, horseradish peroxidase (HRP)-conjugated goat anti-human IgG (diluted 1:5,000, GE Healthcare) was added and incubated for 1 hour at 37°C. The plates were washed again with PBS containing 0.1% Tween 20. Subsequently, 100 μL of HRP substrate (Ncm TMB One; New Cell and Molecular Biotech Co. Ltd, Suzhou, China) was added to each well. After a 15-minute incubation, the reaction was stopped by adding 50 μL of 2 M H SO solution, and the absorbance was measured at 450 nm using a microplate reader. The positive cut-off OD was derived as 3x standard deviation of pre-pandemic sera and was OD 0.5 in this assay.

### Statistical analysis

Characteristics and behaviour of the study participants, stratified by facility type, were tabulated. Chi squared (n≥350) and Fisher’s exact tests (n<350) were used to assess any statistically significant differences in the sex of participants across facility types. A p-value of less than 0.05 was considered statistically significant.

Estimates of viral prevalence and seroprevalence were presented as the number of positive tests over the total number of tests conducted alongside 95% exact binomial confidence intervals, with the latter stratified by both region and facility type.

Due to only one seropositive sample, it was not possible to perform formal statistical analyses of risk factors associated with infection. Instead, a qualitative description of the positive case was included.

Analyses were conducted using Microsoft Excel.

## Results

### Study participants and characteristics

A total of 778 participants were enrolled across the eight study regions and from four facility types (dairy farms, markets, abattoirs and one quarantine station). The samples of eight subjects were spilled during transportation resulting in information from 770 participants included in the final analysis.

Table 1 summarizes the sex, age and duration of work of the study population by facility. Most participants were males (81.6%, 628/770). The proportion of women working in slaughterhouses was approximately twice that of farms (*X^2^*(df=1, N = 609) = 21.86, *p* < 0.001) and markets (*X^2^*(df=1, N = 402) = 14.08, *p* < 0.001), and almost 14 times as great as that in the quarantine facility (Fisher exact test, *p* < 0.001), with the caveat of the substantially smaller size in this facility.

**Table 1.** Demographic profile, duration of work and consumption of camel products as food or

| **Characteristics** | **Slaughterhouse** | **Farm** | **Market** | **Quarantine** |
| --- | --- | --- | --- | --- |
| Total participants enrolled (n) | 288 | 321 | 114 | 47 |
| Female (n (%)) | 84 (29.2%) | 44 (13.7%) | 13 (11.4%) | 1 (2.1%) |
| Age in years (mean (range)) | 37.1 (18-73) | 34.4 (17-80) | 43.6 (18-80) | 34.9 (24-54) |
| Duration of employment in years (mean (range)) | 9.9 (0.6-40)* | 8.5 (0.2-55) | 7.5 (0.2-41) | 7.6 (0.8-13) |
| Used camel product(s) for food (n=593) | 253 (87.8%) | 242 (75.4%) | 70 (61.4%) | 28 (59.6%) |
| Raw milk (n (%)) | 204 (80.6%) | 211 (87.2%) | 56 (80.0%) | 27 (96.4%) |
| Cooked meat (n (%)) | 247 (97.6%) | 226 (93.4%) | 63 (90.0%) | 28 (100.0%) |
| Used camel product(s) for medicinal reasons (n=377) | 115 (39.9%) | 189 (58.9%) | 60 (52.6%) | 13 (27.7%) |
| Milk for constipation (n (%)) | 111 (96.5%) | 152 (80.4%) | 38 (63.3%) | 9 (69.2%) |
| Urine for diabetes & hypertension (n (%)) | 55 (47.8%) | 115 (60.8%) | 30 (50.0%) | 9 (69.2%) |
| Dry stool for skin disease (n (%)) | 49 (42.6%) | 71 (37.6%) | 17 (28.3%) | 2 (15.4%) |
| Blood and stomach content for measles, cellulitis & hepatitis (n (%)) | 19 (16.5%) | 67 (35.4%) | 20 (33.3%) | 1 (7.7%) |
| Bone marrow for fracture (n (%)) | 37 (32.2%) | 73 (38.6%) | 15 (25.0%) | 0 (0.0%) |
| Used PPE items | 66 (16.2%) | 245 (60.2%) | 48 (11.8%) | 48 (11.8%) |

medicinal purposes among study participants, stratified by facility type.
\*Information on employment length at slaughterhouse facilities was available for 284 of 288 participants.

**Table 2.** MERS-CoV seroprevalence among occupationally exposed persons to camels in Somalia, under pseudotype and neutralisation tests. Throughout, we provide the number of positive tests out of the number of total tests, with the corresponding percentage provided in brackets. For aggregated counts per facility or location, we also provide the corresponding 95% exact binomial

| Site/region | Slaughterhouse<br>(n=288) | Farm<br>(n=321) | Market<br>(n=114) | Quarantine<br>(n=47) | Total<br>(n=770) |
| --- | --- | --- | --- | --- | --- |
| **Benadir (n=48)** | 0/5 (0.0%) | 0/34 (0.0%) | 0/9 (0.0%) | NA | 0/48 (0.0% (0.0% - 7.4%)) |
| **Bari (n=84)** | 0/12 (0.0%) | 0/60 (0.0%) | 0/12 (0.0%) | 0/47 (0.0%) | 0/84 (0.0% (0.0% - 4.3%)) |
| **Bay (n=129)** | 0/114 (0.0%) | 0/5 (0.0%) | 0/10 (0.0%) | NA | 0/129 (0.0% (0.0% - 2.8%)) |
| **Galgadud (n=141)** | 0/87 (0.0%) | 0/36 (0.0%) | 0/18 (0.0%) | NA | 0/141 (0.0% (0.0% - 2.6%)) |
| **Karkar (n=84)** | 0/12 (0.0%) | 1/60 (1.7%) | 0/12 (0.0%) | NA | 1/84 (1.2% (0.0% - 6.5%)) |
| **Lower Jubba**<br>**(n=70)** | 0/34 (0.0%) | 0/7 (0.0%) | 0/29 (0.0%) | NA | 0/70 (0.0% (0.0% - 5.1%)) |
| **Mudug (n=83)** | 0/12 (0.0%) | 0/59 (0.0%) | 0/12 (0.0%) | NA | 0/83 (0.0% (0.0% - 4.3%)) |
| **Nugal (n=84)** | 0/12 (0.0%) | 0/60 (0.0%) | 0/12 (0.0%) | NA | 0/84 (0.0% (0.0% - 4.3%)) |
| **Total (n=770)** | 0/228<br>(0.0% (0.0% – 1.6%)) | 1/321<br>(0.3% (0.0% – 1.7%)) | 0/114<br>(0.0% (0.0% – 3.2%)) | 0/47<br>(0.0% (0.0% - 7.5%)) | 1/770<br>(0.1% (0.0% - 0.7%)) |

### Consumption of camel products

Table 1 presents the summary of regular use of camel products for food or medicinal purposes by participants. Of 770 respondents, 593 (77.0%) reported regularly consuming camel products for food, primarily raw milk (84.0%; 498/593) and cooked meat (95.1%; 564/593). None of the respondents reported consuming raw meat or urine for food. Almost half (49.0%; 377/770) of the respondents stated that they use camel products for medicinal purposes. Specifically, 82.2% (310/377) reported drinking raw milk to treat constipation, 54.4% (209/377) reported drinking urine to treat diabetes and hypertension, 36.9% (139/377) reported using dry stool to treat skin diseases, 28.4% (107/377) reported smearing blood and stomach contents on the skin to treat measles, cellulitis and hepatitis and 33.2% (125/377) consuming bone marrow to heal fractures. Study participants from farms were significantly more likely to report using camel products for medicinal products compared to slaughterhouses and the quarantine facility (farms: 58.8%; slaughterhouses: 39.9%; *X^2^* (df=1, N = 609) = 6.98, p=0.008; quarantine: 27.7%; Fisher exact test, p=0.022), but not compared to participants in markets (48.6%; *X^2^* (df=1, N = 609) = 1.06, p=0.304).

### Use of Personal Protective Equipment (PPE)

The different PPE items used by various roles of workers and did not show any differences for those who had used PPE or not during activities with animals. However, those with animal contact in their role claimed to use more PPE than those with work not associated with animal care. Of those who claimed general use of PPE during activities (33.4%; 257/770), the majority reported just glove use (58.0%; 149/257). 11.7% (30/257) of respondents reported using gloves/masks, 10% (25/257) reported using only boots, and 3% (8/257) reported using only masks, with the rest (16%; 41/257) reporting the use of a combination of coveralls, masks, gloves and/or boots.

### Detection of MERS-CoV

None of the naso/oral pharyngeal swab specimens were positive for MERS-CoV RNA by RT-PCR (0.0% (95% CI 0.0% - 0.5%)).

A total of 18 samples had positive or borderline positive results for anti-MERS-CoV IgG when tested at the local laboratory using ELISA (2.3% (95% CI 1.4% - 3.7%)). All 18 positive samples and 73 randomly-selected negative samples were tested by EUROIMMUN MERS-CoV human IgG ELISA and a pseudoparticle neutralization test at the University of Hong Kong laboratory. One serum was IgG positive and two were borderline positive. The ELISA positive sample was also confirmed to be positive in MERS-CoV IgG pseudoparticle neutralization tests with a titre of 1:10. This serum sample and 34 other sera including those with borerline or positive ELISA results in the field laboratory were tested in MERS-CoV PRNT tests. The serum sample positive in pseudoparticle neutralization was the only serum positive in PRNT, with PRNT50 titre of 1:40 and PRNT90 titre of 1:20. This finding constitutes the first evidence of zoonotic spillover of MERS-CoV in Somalia with an overall seroprevalence estimate of 0.1% (95% CI 0.0% - 0.7%).

The seropositive case belonged to a male in the 30 – 39 years old age group who worked full-time for several (5 – 10) years as a livestock handler on a mixed-use farm in Qardo district, Karkar region, Puntland. The individual was responsible for daily animal care activities, including feeding, assisting in birthing, administering veterinary treatments, and directly handling camels, goats, and cattle. His role included milking camels and cleaning animal enclosures, which placed him in frequent, close contact with animal bodily fluids and waste. The individual had no known pre-existing medical conditions and reported good general health. He reported recent travel to the regional livestock market two weeks before sampling, where he engaged in trading activities and had close contact with traders and livestock from different regions. However, at the time of sampling he reported no international travel at any point in his life, indicating the first evidence of MERS-CoV spillover infection into humans in the country.

The seropositive case reported irregular use of protective gloves and no use of masks or protective outerwear during animal handling tasks. Hand hygiene was reportedly infrequent after contact with animals or animal products. The individual regularly consumed unpasteurized camel milk and occasionally ate raw camel liver. No family member reported symptoms at the time of the interview.

Ninety one serum samples were tested for SARS-CoV-2 IgG antibodies and 81 (89.0% (95% CI 80.8% - 94.6%) were positive, including the MERS-CoV seropositive sample.

## Discussion

Evidence of past MERS-CoV infection was confirmed with neutralisation testing in one individual occupationally exposed to camels in Qardo district, Karkar region, Puntland, Somalia. As this serum sample was collected in an individual who had never travelled outside the country, this finding is the first evidence of MERS-CoV zoonotic spillover to occupationally exposed humans within Somalia. The positive case, a male livestock handler on a mixed-use farm aged 30 – 39 years old, was responsible for daily animal care activities including milking camels and cleaning animal enclosures, which placed him in frequent, close contact with animal bodily fluids and waste. Notably, he reported recent travel to the regional livestock market in Somalia where he engaged in trading activities and had close contact with traders and livestock from different regions. The individual reported irregular use of PPE or hand sanitation, regular consumption of unpasteurized camel milk and occasional consumption of raw camel liver, both products recognized for their risk of exposing people to infection when consumed raw. No other family members reported symptoms at the time of the interview, and household-level investigations did not reveal additional cases, suggesting no evidence of onwards transmission.

As serology indicates evidence of past infection, it is not unexpected that this individual was negative for virus RNA in RT-PCR tests. Given that MERS-CoV virus RNA detection in the respiratory tract is short lived (Park et al., 2018), the window of detection for such infection is much shorter than with serology. There is only one report of detection of MERS-CoV RNA (by RT-PCR) in humans in Africa and that was in a study of camel herders in Kenya, where MERS-CoV was detected in 3 of 262 camel handlers (Munyua et al., 2019). Therefore, it is important to carry out larger studies of camel-exposed individuals in Somalia to be able to detect serological responses and understand the extent of transmission in the population. In our study, the absence of concurrent sampling of camels limits direct inference about animal-to-human transmission dynamics at the time of exposure.

Serological evidence of MERS in humans has also been reported in Kenya and Morocco (Liljander et al., 2016, Munyua et al., 2019, Abbad et al., 2019). In addition, studies of patients with symptomatic disease confirmed by RT-PCR have shown that more severely ill individuals invariably undergo seroconversion, as detected by IgG ELISA and neutralization tests, and that these antibody responses persist for at least four years before gradually waning (Cheon et al., 2022). In contrast, MERS-CoV seroconversion occurs only in some mildly ill or asymptomatic individuals, with others remaining sero-negative even through having RT-PCR confirmed MERS disease (Choe et al., 2017) (Kim et al., 2024). Serology is therefore likely to underestimate the true extent of MERS-CoV infection in a population. However, some RT-PCR confirmed individuals appear to mount MERS-CoV specific T cell responses (Zhao et al., 2018). Camel abattoir workers in Kano, Nigeria were MERS CoV sero-negative, but one third of them had detectable MERS-CoV specific T cell responses (Mok et al., 2021). Thus, future studies could investigate MERS-CoV specific T-cell responses in camel-exposed human populations in Somalia and elsewhere in Africa, to better define the true extent of human infection taking place in the region.

Although 89% of the sub-sample of sera tested for SARS-CoV-2 were seropositive, indicating past COVID-19 infection or vaccination, the EUROIMMUN MERS-CoV IgG ELISA remained highly specific when carried out in the reference laboratory in the University of Hong Kong. These findings support the reliability and specificity of the EUROIMMUN MERS-CoV IgG ELISA even after the worldwide spread of SARS-CoV-2. Although 18 serum samples tested MERS-CoV IgG borderline positive or positive in the field, only one of them could be confirmed by IgG ELISA in the reference laboratory and this serum was also confirmed positive in pseudovirus neutralization and PRNT assays. The false positives generated when testing the samples with the EUROIMMUN MERS-CoV IgG ELISA in the field may be due to challenges in performing a new assay correctly and should be followed up through targeted training.

While MERS human disease to date have been reported only from or with an exposure link to the Arabian Peninsula, field research has shown that MERS-CoV is circulating widely in camel populations in several African countries. MERS-CoV in African camels belongs to clade C while those causing human disease in the Arabian Peninsula have been clade A or B MERS-CoV (Ware H*, 2024) (WHO,2025). Clade C MERS-CoV viruses appear to have lower replication competence in the human respiratory tract (Zhou et al 2021). However, there are studies producing serological, T cell response and RT-PCR data that suggest zoonotic spillovers are taking place in the continent (Liljander et al., 2016; Abbad et al., 2019; Kiyong’a et al., 2020). Although the serology assay cannot distinguish between infecting clades of MERS-CoV, these findings in Kenya, Morocco, Algeria and now Somalia, indicate that clade C MERS-CoV can infect humans and that such infections are occurring within camel-herding regions of Africa. It is noteworthy that Clade B MERS-CoV has recently been reported in camel populations in Egypt and Sudan (Gomaa et al., 2025), highlighting the risk of introduction from the Arabian Peninsula into African camel populations and further spread in light of its increased replication competence and more efficient viral entry over Clade C. (Haroun et al., 2023)

Africa hosts 75% of the global dromedary camel populations, and it is unsurprising that MERS-CoV circulates in African camels as it does in camels in the Arabian Peninsula (Miguel et al 2017; Sikkema et al 2019). High-risk human activities, such as farming, that increase exposure to dromedary camels are as common in Africa as in the Arabian Peninsula (Abbad et al 2018; So et al 2016), providing ample opportunity for zoonotic infection to occur in the human population, as evidenced by recent sero-epidemiological studies including this one. Despite these risks, at present, there is little awareness and testing of MERS-CoV as a cause of respiratory disease in Somalia and the African continent more broadly. The viral respiratory sentinel surveillance that is taking place, such as influenza-like illness (ILI), severe acute respiratory infection (SARI) surveillance and integrated disease surveillance and response (IDSR) sites are not necessarily located in camel-herding populations. MERS-CoV has been recognized by WHO as an epidemic- and pandemic-prone pathogen regardless of its geographic origin, highlighting the need to enhance awareness and surveillance for MERS-CoV in camel herding regions across Africa.

## Conclusion

Our study provides the first-ever evidence of zoonotic spillover of MERS-CoV to humans in Somalia and adds to accumulating evidence of zoonotic spillover of MERS-CoV in Africa. These findings underscore the need to recognize MERS-CoV as a potential aetiological agent of respiratory disease within Africa. The findings emphasize the necessity of enhancing surveillance and monitoring systems for MERS-CoV in Somalia, alongside strengthening national and state capacity to produce accurate and reliable MERS-CoV data. Further studies on MERS-CoV infection in camels as well as understanding the socio-cultural risk factors will be crucial for developing effective and targeted prevention and control strategies.

## Authors’ contribution

MW, JA, ML, MMB and SvD designed the study. MW, JA, AS, MP, RM and SvD contributed to the analysis of the data and wrote the first draft of the manuscript. MMH and SIM coordinated blood sample collection and conducted in-country laboratory analysis. MMH, SIM, AAA and BAJ designed the database and contributed to data validation and cleaning. MMH, SIM, AOA, AbMA, AYA, AhMA, AdMA, OAA, AMM, AAB, AJF, AMHH, AMH, MoAH, MuAH, FAH, HAH, MoAM, MuAM, OMM, MBN, OAS, AAbS and ANFS contributed to the study implementation, data collection and data entry online. SMSC, JCHH, JKCL and MP undertook the validation and confirmatory analysis of the blood samples. HAEN, AAlS, MDVK, AB provided technical guidance for the study. AS, ML, RM and SvD provided project management and administration related to the paper. All authors contributed to redrafting and approved the final manuscript.

## Disclaimer

The views expressed in this article are those of the authors and do not necessarily reflect the official policy or position of WHO, or any other affiliated institutions.

## Financial support

The study was supported by WHO and funded by GIZ One Health (grant number 81278362). RM acknowledges funding from the Medical Research Council (MRC) Centre for Global Infectious Disease Analysis (MR/X020258/1) funded by the UK MRC and carried out in the frame of the Global Health EDCTP3 Joint Undertaking supported by the EU.

## Data sharing

Data may be available upon reasonable request to the corresponding authors.

## Conflicts of interests

No authors have reported a conflict of interest.

## Data Availability

Data may be available upon reasonable request to the corresponding authors.

## Acknowledgement

We would like to thank study participants and authorities of the study facilities for their cooperation and support.

## Notes

### Competing Interest Statement

The authors have declared no competing interest.

### Author Declarations

Ethical Clearance was approved from the East Africa University Research Ethical Review Committee.

